# A qualitative study of anaesthetic trainee and trainer perspectives on regional teaching programmes

**DOI:** 10.1101/2024.01.03.24300775

**Authors:** A Lee, R Clementi, S Jackman, M Rakic, S Smyth, R Wight

## Abstract

**Summary:** *Background:* Regional teaching programmes are a common method of delivering postgraduate medical education in the United Kingdom. There is limited evidence available to guide the development of a successful regional teaching programme.

*Methods:* In this qualitative study, we interviewed stakeholders including trainers and trainee anaesthetists using a combination of purposive and convenience sampling. Semi-structured interviews were conducted on Microsoft Teams in a focus group format. Data was transcribed, anonymised, coded and analysed using thematic analysis.

*Findings:* All the college tutors and school board members for anaesthesia in the deanery, and five anaesthetic trainees at different training stages participated in the study in six focus groups. Analysis revealed concerns about the complicated nature of Stage 1 anaesthetic training and discussion about how the local training structure within the deanery could be further impeding the delivery of support and education to Stage 1 anaesthetic trainees. Despite the challenges identified, trainers and trainees were optimistic about implementing a regional teaching programme for Stage 1 anaesthetists within the deanery.

*Conclusions:* This study highlighted the challenges of delivering high-quality postgraduate education and training for Stage 1 anaesthetists in our deanery and considered possible solutions. The findings of this study informed the development of a quality improvement project to improve Stage 1 anaesthetic training in our deanery.

## Introduction

Regional teaching programmes are a common way of delivering postgraduate medical education in the United Kingdom (UK). The rotational model of training in the UK necessitates a way to deliver standardised, specialty specific teaching for doctors working across multiple sites. The General Medical Council (GMC) includes a question about the quality of regional teaching in the annual National Training Survey[1].

There is no guidance on how to deliver a regional teaching programme for any level or specialty in the UK, although such programmes are very common. There is limited published data [2,3] on regional teaching programmes in the UK, likely in part due to the complexity of providing guidance on designing regional teaching programmes for trainees with differing educational requirements and the wide range of geographical distributions of regions of training (which can differ between specialties)[4].

Our deanery provides Stage 1 or core anaesthetic training across five hospital trusts. There are approximately 50 core anaesthetic (3 years, CT1-3) or acute care common stem (ACCS) anaesthetic trainees (4 years, CT1-4) across the deanery.

The national 2021 change in curriculum for anaesthetists [6] prompted a review of Stage 1 anaesthetic training provisions within the deanery. At the time, no regional teaching programme for Stage 1 training existed within the deanery, and 5-year GMC survey results demonstrated low scores for regional teaching for several hospital trusts[1].

A quality improvement project (QIP) was launched to understand the situation regarding the current teaching provision for Stage 1 anaesthetic trainees in the deanery, investigate whether improvements were needed and to implement change where necessary.

This paper describes the qualitative research component of the QIP, where trainees and trainers in the deanery were interviewed to understand their perspectives on regional teaching for Stage 1 anaesthetists in the deanery.

## Methods

### Approach

We chose a qualitative approach because it can capture complex ideas that cannot easily be derived from quantitative surveys.[7] We were conscious of the lack of existing evidence and guidance on regional teaching programmes and wanted to understand the meaning that stakeholders of the situation attributed to it before making any hypotheses or implement any changes. Thematic analysis was chosen as the analytic method due to its flexibility to be applied in an inductive manner for this dataset, generating new themes based on patterns within the data.[5]

### Procedure

We performed purposive sampling of all core training college tutors and anaesthetic school board members in the deanery. We performed convenience sampling for trainees via e-mailing lists and trainee social media groups invitations for a 4-week period and closed recruitment after no new participants came forward for more than 7 days. All participants were given written information by email about the study procedures and gave written or verbal consent to participate. At the start of each interview, participant consent was reconfirmed by Author1, including permission to record and transcribe the interviews. We grouped participants into focus groups based on availability, as all interviewees were full time clinicians with busy schedules at the time of interviews.

Author1 had met or worked clinically with most of the trainees and trainers interviewed for the study. All interviewees were given written information by email about Author1’s role and the purpose of the study, and this was reconfirmed by Author1 at the start of each interview. The other authors received informal training from Author1 in coding and qualitative methods.

All interviews were semi-structured and recorded in audio and video form with a reference automated transcript generated by Microsoft Teams. All recordings were manually transcribed and anonymised by the research team. Transcripts were not returned to participants, but all participants reviewed the draft of this manuscript before manuscript submission.

Interview questions had been pilot tested between Author1 and the research team prior to the first focus group interview and refined accordingly. Thematic analysis was performed on the corrected transcriptions as follows: 1) individual researchers coded data and identified relevant themes; 2) the research group met to discuss overarching themes; 3) the data was revisited, and the final set of overarching themes were refined by Author1.

The six researchers who performed coding recorded field notes pre- and post-interview and after individual coding regarding their own perceptions and biases to ensure reflexivity. Data was coded using Microsoft Word or on paper. A completed Consolidated Criteria for Reporting Qualitative Research (COREQ) checklist [8] is provided in the appendix.

### Findings

Six trainees and eight trainers were interviewed separately in focus groups of 2-4 participants each on Microsoft Teams by Author1. Table 1 shows a list of focus groups conducted and participants in each group.

**Table 1:**

| Focus groups | Participants |
| --- | --- |
| Group 1 | College tutor 1<br>College tutor 2 |
| Group 2 | College tutor 3<br>College tutor 4 |
| Group 3 | College tutor 5<br>School board member 1<br>School board member 2 |
| Group 4 | School board member 3 |
| Group 5 | Trainee 1 (Stage 1)<br>Trainee 2 (Stage 1)<br>Trainee 3 (Stage 2)<br>Trainee 4 (Stage 1) |
| Group 6 | Trainee 5 (Stage 1)<br>Trainee 6 (Stage 2) |

Each interview lasted between 40 minutes to 1 hour. There were no dropouts and all trainers participated in the study. All trainees worked within the deanery as anaesthetists at the time of the study, and included 2 registrars, ST4 and 5 respectively (one who had completed core training in the deanery and another who had completed core training in a different deanery), and 4 Stage 1 trainees from CT1-3. All five college tutors from each of the hospital trusts in the deanery, the head of school and training programme directors for core and registrar training participated in the interviews.

Figure 1 shows the code tree for the themes derived from the data.

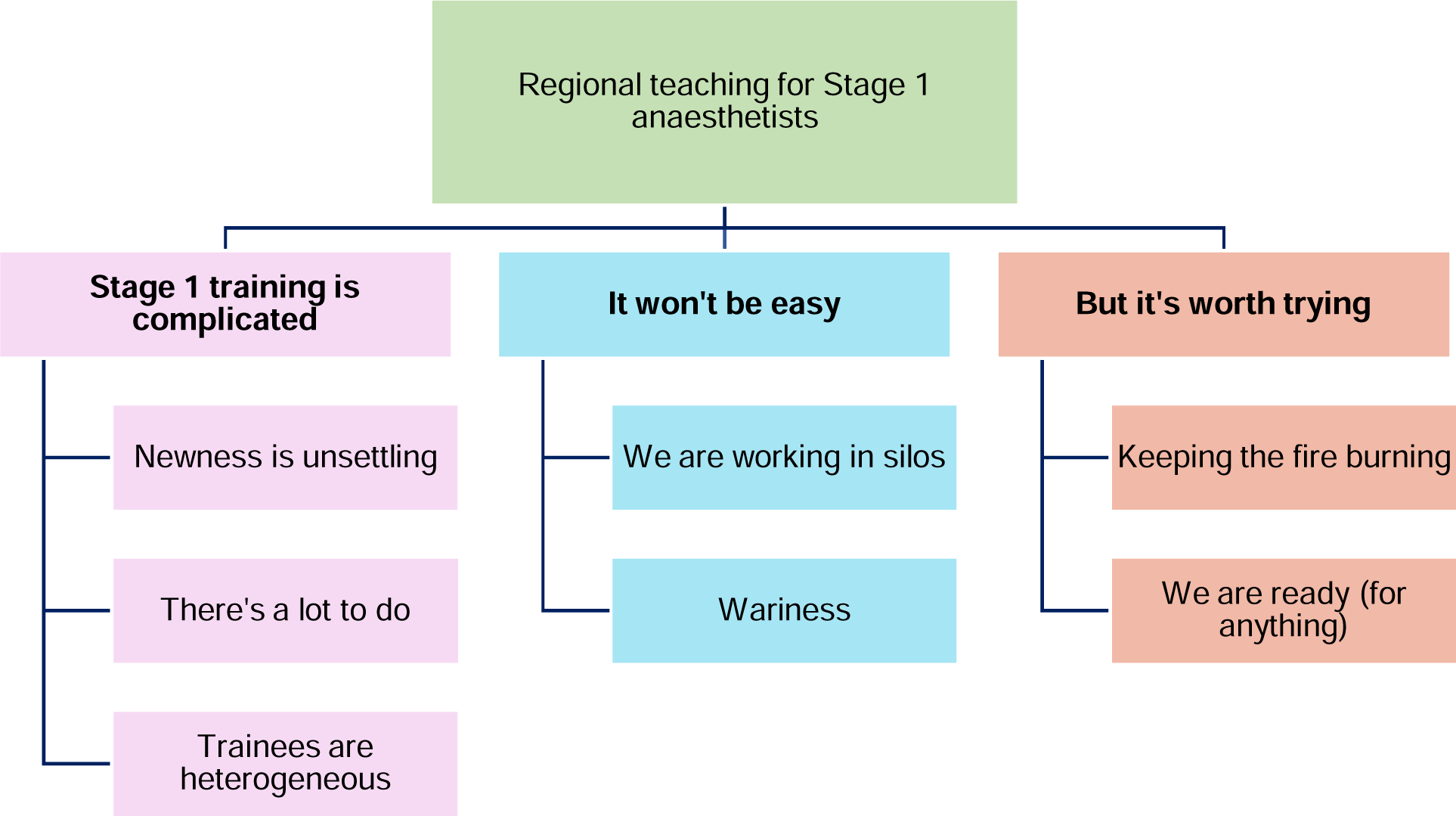

### Stage 1 training is complicated

#### Newness is unsettling

The new stage 1 curriculum had introduced an extra year of training to Stage 1 anaesthetists, with some additional clinical learning needs and an ongoing existing requirement that the three-part primary Fellowship of the Royal College of Anaesthetists (FRCA) examination was passed by the end of the programme.[6] This led to many changes within the deanery, with Hospital 2 having stage 1 trainees for the first time, and a mixture of stage 1 trainees on the old or new programmes in various hospitals with different learning needs. Trainers and trainees identified that these changes led to a state of uncertainty, with unclear expectations between both groups.

> College tutor 2

> “I’m glad that the training has shifted and the program has changed, but… we haven’t had any true stage 1 trainees start and go through the program yet. They’re, we’re all in a state of flux..it’s going to take a while to, to sort of settle that down..”

> Core trainee 5

> “..it’s the first year this has happened.. and it’s a bit of a surprise for everyone and it’s gonna take a little bit of time to work out exactly.. what is expected of us in terms of .. responsibilities and experience.”

### There’s a lot to do

All trainers discussed how they all felt there was a lot to achieve in the 3 years of Stage 1 training (“I think it’s amazing that people ever got through in two years” College Tutor 2). Several pointed out how job applications for stage 2 (registrar) training occurred approximately 30 months into the Stage 1 programme, after which it was no longer possible to add curriculum vitae (CV) items or exam results into an application to Stage 2 to follow directly after Stage 1.

While this was an improvement on the original 18 months, trainees are still expected to complete Stage 1 with good clinical competence, passing the primary FRCA examination, and CV and extracurricular development that would make them competitive at a Stage 2 job interview. Trainers were sympathetic about how challenging this was for trainees (‘I see that it’s a very stressful time..’ College tutor 5). Trainers described feeling responsible for all these aspects for their Stage 1 trainees, and wanting them to enjoy their training despite these high pressures (‘..if [trainees] are happy, I’m happy’ College Tutor 3).

> College tutor 1

> “..in that time they have to complete the primary examination before they can move on to specialist training and the competitiveness of getting onto a training program has gone through the roof.. there’s a big step up for what people actually need to achieve in that period to actually get themselves into an ST program.”

> College tutor 2

> “..it’s taking people from a place of innocence, so, unable… that’s a bit of a strong word, but… not, not able to give an anaesthetic, knowing very little, if anything, of how to do anaesthetics, to fairly swiftly being actually quite independent practitioners. So there’s a big clinical demand on them, particularly in the first six months to a year..to be able to give straightforward anaesthesia alone.”

The primary FRCA exam was mentioned repeatedly as a major milestone for Stage 1 training. Trainees often referred to the support given to them for exam preparation as a key measure of their satisfaction with their Stage 1 training.

> Trainee 3

> “I had a really great experience of core training in this deanery. I think, particularly where I did CT1, the exam teaching there was excellent..”

Both trainees and trainers considered exam preparation as a separate entity to clinical experience and CV building opportunities. It was evident that neither party particularly enjoyed teaching nor learning for the exam, a situation mainly attributed to the heavy scientific content in the primary FRCA exam. Trainees also commented that some of the scientific teaching was of low quality, possibly as a result.

> “..hardcore painful lectures..” College tutor 2 “..soul destroying..” College tutor 1

> “Consultants do not like to teach primary FRCA because of the basic sciences” School board member 3

> “.. pure primary drudgery, everyone would fall asleep and be hating life by the end of it” Trainee 3

> “..spectacularly badly taught…blind leading the blind” Trainee 4

Several trainers were concerned that the increase in duration of training would not solve the original problem where trainees were unable to complete all their stage 1 requirements at the end of the programme. The primary FRCA had often been the limiting factor for this for many trainees in the past. Trainers were concerned that trainees might delay starting to sit the primary FRCA and still not complete the exam by the end of Stage 1, despite having the extra year.

> College tutor 1

> “There could be – a mission creep where everything’s just done later and you still have the same problem that trainees are currently facing: instead of it being at the end of their second year, it’s at the end of their third year.”

### Trainees are heterogeneous

Stage 1 trainees were described as a particularly heterogeneous group in terms of learning needs, experience, and training requirements (“they want different things at different times” College tutor 2). Trainers felt that there was no one size fits all model of training that could be used for all trainees as Stage 1 trainees ranged from those newly starting anaesthetics to trainees able to independently run anaesthetic services for busy maternity units or theatre suites overnight.

> School board member 2

> “..trainees lie along the spectrum..the ones who embrace the hurdles and get on with them quickly because they see that as what they need to do and projects and exams et cetera and the ones who want to be told what to do a little bit more and have very specific direction.”

Trainees themselves also recognised the differences between them, and how this made it challenging to structure and deliver appropriate teaching for their varying educational needs.

> Trainee 2

> “..hopefully like there could be teaching focused on people who are preparing for the primary. There could be teaching focused on people who are preparing for the final and then there will be people who aren’t really focusing on an exam at the time..”

This issue of trainee variety led to a lot of discussion between trainees and trainers about many teaching sessions being organised for Stage 1 trainees but ending up missing the mark with content or audience, leading to dissatisfaction.

> Trainee 2

> “..some trusts, the teaching is for core level is merged with non-career anaesthetists.. can be quite vague and not very exam focused which when you’re coming up to an exam is more what you want.

> ..another trust ran most of its teaching monthly in terms of big sims in terms of sim days which again was it was fun and engaging, and useful practically but no use really for the exam except for maybe if you were fortunate enough to get one OSCE station on a sim you’ve done before.”

Trainers agreed that the combination of the new curriculum, high demands of training and wide range of trainee needs and abilities had spurred them on to consider new ideas to deliver training and support the Stage 1 trainees in the deanery. These concerns and ideas such as a unified teaching programme were raised repeatedly during school board meetings in the preceding 12 months and eventually reached a “tipping point.. rather than saying it’s a good idea, [we wanted] to actually push forward and start doing it” (School board member 1).

### It won’t be easy

#### Working in silos

All interviewees described a significant variation in how each of the deanery’s hospitals operate and provide anaesthetic training. The term “fairness” came up repeatedly during discussions about the differences between sites for all interviewees, with many feeling that the current system was an unfair lottery that was doing the trainees a disservice.

Trainers were ‘aware’ that other hospitals had different teaching offerings but without significant detail, and trainees would rotate to different hospitals each year to join a different programme, with a risk of “missing out on certain little chunks of the curriculum” (College tutor 3). Trainers at hospitals 1,4 and 5 who had few career anaesthetist trainees described great difficulties organising teaching for their smaller number of trainees.

> College tutor 4

> “quite difficult because you’ve got trainees at different stages of training with different exams sitting in on the same [teaching session].. core training specifically is probably a little bit lacking [in this hospital]”

> College tutor 1

> “they’re going through this process alone as an individual with a teaching program undertaken within the hospital that’s not bespoke for their learning requirements… we are inhibiting the trainees coming through [this hospital]”

> College tutor 5

> “we find it still, difficult to run a full-fledged educational program mainly because..the smaller numbers that are there and which makes it..difficult to make it viable..”

Trainees described frustrating experiences with variable teaching as they rotated through hospitals and worry that they might be in a hospital with poorer exam support at the time of their exams.

> Trainee 3

> “I was at Hosp3 and I think their teaching is exceptional.. other hospitals I’ve been to the teaching just does not compare.. unfair towards trainees who do placements at those hospitals at the time that they’re doing their exam.”

> Trainee 5

> “massive variations from hospital to hospital..unfair discrimination depending on where a trainee might be working..poor quality educational experience”

> Trainee 4

> “[trainees in hospital 3] probably don’t know how good they’ve got it”

The differences between training offerings between hospitals fostered a culture of unhealthy competition between them, a situation picked up by the trainers and trainees alike. Trainers at the hospital that had the highest regarded teaching (hospital 3) felt disincentivised from sharing their expertise with the others, while the other hospitals suffered from poor trainee feedback because they were often unable to provide the same quality of teaching. It was generally agreed that one of the main reasons for hospital 3’s success was the fact that they only had career anaesthetists training there and had the highest number of career anaesthetists at stage 1 of all the sites, making it easier to provide high quality exam-focused teaching.

> Trainee 2

> “Hospital 1..has a high opinion of itself because it is well run. And I think it is a little bit peeved by how much the trainees rotating rave about Hospital 3’s teaching.”

> College tutor 3

> “[Hospital 3] probably got the most developed teaching in the region. we’re giving up the jewel in the crown [by participating in a regional programme]”

> School board member 1

> “Concerns..from Hospital 3 on occasion as to whether they could quality assure that training if..they would have to stop delivering their in-house teaching [by participating in a regional programme]”

Hospitals providing different compensation to trainers for teaching was also a point of tension between the hospital sites in the region. Some hospital trusts offer consultants paid teaching time and others do not. School board members pointed out that this was going to be an ongoing sticking point if a regional teaching programme was being considered, as funding for postgraduate education comes from individual trusts instead of the deanery or school of anaesthesia. College tutors described their role as complex and often consuming more time than allocated (“we’re all on one PA to look after 25 trainees each” College tutor 2), leaving little time to devote to organising teaching in addition to their many responsibilities.

> School board member 1

> “if you’re in one trust and..you’re a keen educator..we’ll give you a PA to go [and teach]. And you’re in another trust..we’ll give you no money but you’re very welcome to go.”

> College tutor 2

> “I could get another half PA to coordinate teaching, but I’d have to give up more clinical”

#### Wariness

Not all the interviewees felt that a regional teaching programme would be a magic solution to solve to the challenges already identified: the complex Stage 1 training programme and disparities between hospital sites in the region. College tutor 3 spoke about ‘mixed feelings’ and having to consider why a programme had not been previously implemented. Trainee 3 felt that a poorly executed programme would be a “bad idea.. probably worse than what we have already”. School board member 3 had particularly different views from the other interviewees. They had previously discussed regional teaching with a few trainees and felt that trainees had “no appetite” for regional teaching, preferring local teaching instead (“I have yet to find a single trainee who wants [regional teaching]”); but all the trainees interviewed thought a good regional teaching programme would be an asset to their training. These differing views highlighted the gaps in our understanding about regional teaching in the deanery, and clearly demonstrated that tools such as the GMC survey are insufficient to give trainers enough information about what trainees want, or what the next steps should be.

There were several other logistical challenges identified for implementation of a successful programme, with participant quotes shown in Figure 1. Location and mode of teaching was a key challenge discussed in all the interviews, as a regional programme would mean getting trainees together from different sites across the region. Several hospitals were particularly short of teaching spaces for higher numbers or had limited availability for rooms. When considering format, some worried that trainees might not be keen to attend a fully face-to-face programme due to difficulties with transport arrangements or that face-to-face sessions might discriminate against trainees who were unable to drive. These concerns had to be weighed against the reduced social aspect, interactivity, and engagement potential of a fully online programme; or the complexities of organising a hybrid programme.

There were comments that a bigger group might change the dynamic of a teaching session, making it less easy to keep a session interactive or engaging. Rota organisation for most sites would be more complicated, as trainees on an evening shift might face a long commute back to their base hospital to attend an evening shift or miss the teaching day completely; and less than full time trainees might have to miss sessions on their non-working days.

The increased travel requirements may lead to less frequent full day instead of more frequent half-day sessions, but this might mean that trainees missing a session would miss a higher proportion of overall teaching. Funding for travel or rooms was another significant concern, as the deanery did not offer additional funding to cover the costs of regional teaching programmes. Trainees could potentially claim travel expenses from their study budget but only after a certain distance had been incurred, and it was unclear if claiming might affect their ability to attend other exam related courses using their study budget.

Quality control for teaching was another important aspect to consider: all trainers and trainees felt that the challenges described were not insurmountable, but that the teaching had to be of high enough quality to merit the effort it would take from all parties to overcome them. A programme would also have to maintain its high standards moving forward, which would require active monitoring of session feedback and a willingness to adapt sessions for the future.

**Figure 1:**
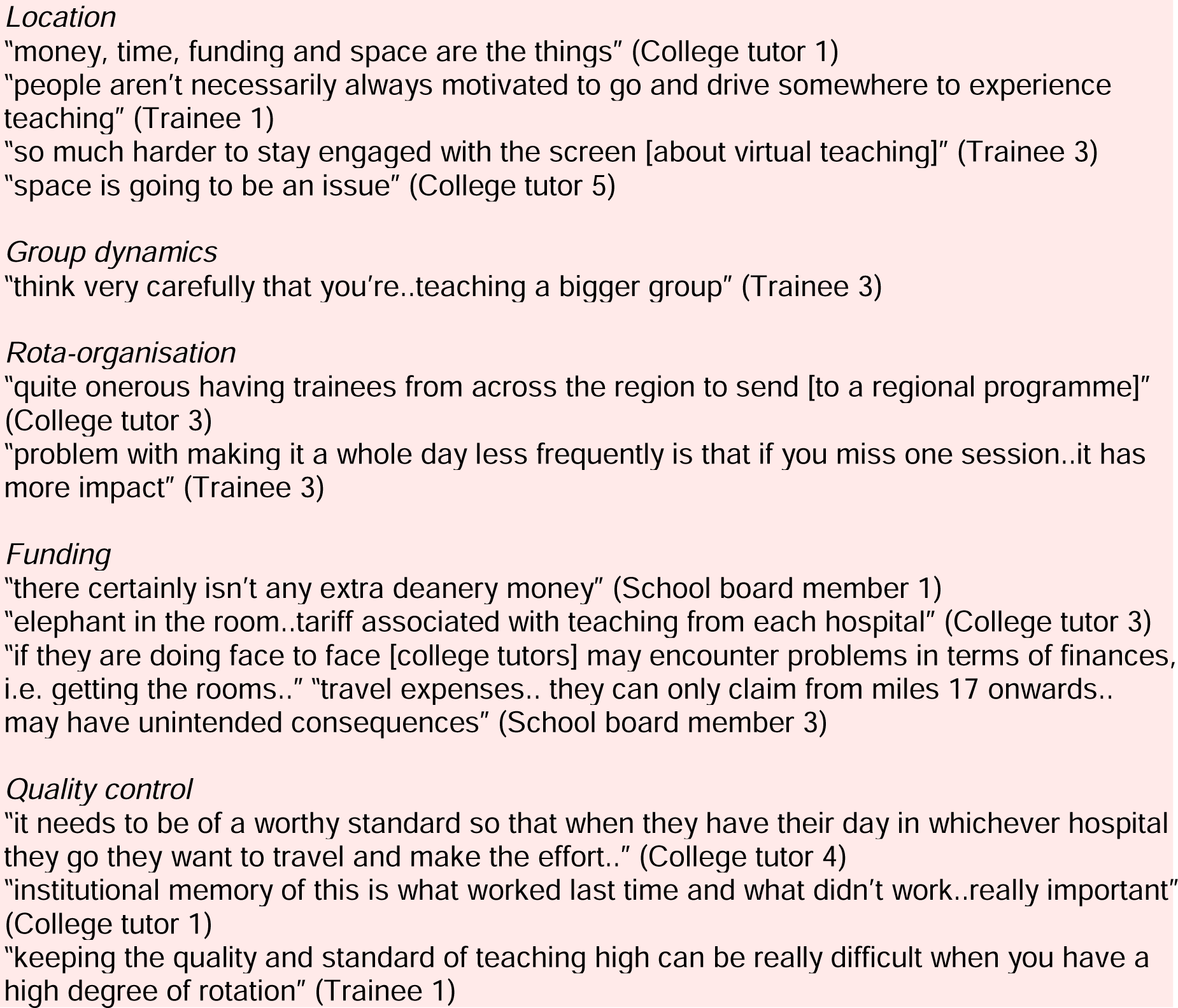
Logistical challenges associated with a regional teaching programme

### But it’s worth trying

#### Keeping the fire burning

All interviewees identified positive motivators to launch a regional teaching programme in the deanery. Participant quotes are shown in Figure 2. All the trainers were enthusiastic about being able to offer a lot more to the trainees with a regional programme, with better exam-focused teaching and skills including wellbeing and human factors. There were many ideas about developing a body of ‘Stage 1 or primary FRCA’ focused educators or sessions for the course and giving trainees the benefit of the best teaching that each centre could offer. This could come with many benefits, overcoming challenges of quality control and consistency. Many felt that a programme would help standardise the knowledge and exposure for all the Stage 1 trainees, improving fairness. Smaller hospitals would find it easier to join a bigger group and share training resources, and trainees in smaller hospitals would not feel as isolated. Trainees felt it was a great opportunity for them to meet their colleagues and that this might help with increasing cross-linking between hospitals, both clinically and with other projects.

**Figure 2:**
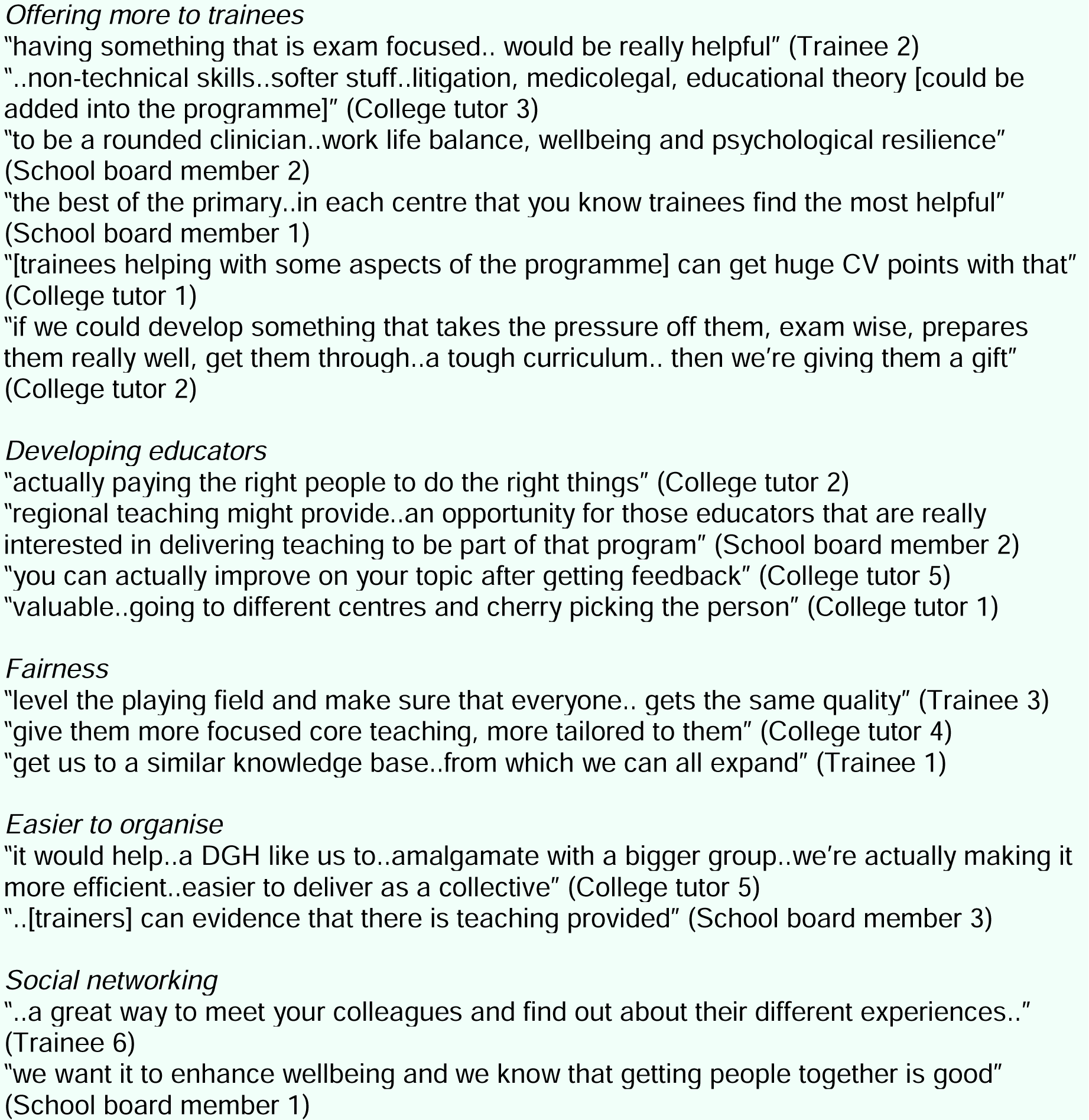
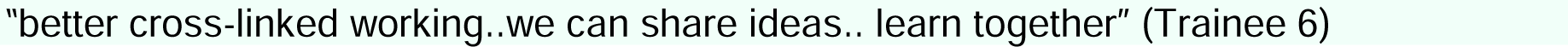
Motivators to launch a regional teaching programme

When discussing the sustainability of the programme, most interviewees felt that a high-quality programme would sustain itself. There had been precedence in the region that two anaesthetic courses had been developed and launched by enthusiastic trainees and had become part of the organisational culture. Both courses continue to run at a high standard and are run by trainees on a volunteer basis (unpaid, without specific time allocated within their jobs). Most interviewees mentioned both courses as references for success and considered that the reasons for success included high-quality teaching that was of interest to trainees or specific teaching for a specific group of learners unavailable elsewhere, and several generations of interested coordinators who had spurred the courses forward.

Trainees referenced the deanery’s acute care common stem (ACCS) regional teaching course as an example of a poorer precedent within the region, as it is a trainee run course without a specified overall coordinator, providing teaching to emergency medicine, intensive care, acute medicine and anaesthetic ACCS trainees (“I think it was basically just handed to the trainees and said go ahead, organise it yourself” Trainee 2). Although it continues running, the 3 ACCS trainees in the interviews unanimously felt that it could be improved. They identified having one rotating trainee in charge of a teaching day; a disparate audience with different learning needs; and lack of resource or training to organise teaching as the main reasons for its lower quality. They felt that a regional teaching programme that was primarily focused on the primary FRCA exam (same learning needs) for Stage 1 anaesthetists (same type of audience), supervised and run by senior trainees or consultants (experienced and identified coordinators) would be far more likely to succeed.

#### We are ready for anything

All trainers felt ready to embrace a new programme if the quality improvement project revealed that this was something that the trainees wanted. All the college tutors felt that there were more positives than negatives in having a programme and spoke about working together between them to help make the programme work.

> College tutor 3

> “I think combining across the region means that.. there’s a sort of..an impetus.. like a momentum with that and that’s probably more exciting.”

> College tutor 4

> “If this does happen, then we’re all going to be collaborating together to do it..we’re all on board and committed”

> College tutor 5

> “it’s in everybody’s interest to keep this a success and keep it going.. it’s a good collective effort by multiple different hospitals..more chances of this succeeding than a local teaching programme”

When discussing a potential new programme, all trainers had ideas for how an ideal programme should run but recognised that the quality improvement project might reveal that trainees had different ideas. They felt that the trainees’ opinion took precedence over their own when it came to programme design and were willing to try whatever was suggested.

> College tutor 4

> “[a new programme] might be change that works very well. It might be change that doesn’t work, but you’ve got to do it to know if that makes sense.”

> College tutor 2:

> “You know we support you and I think what we have to be respectful of is that you’ve come out with the program we need to..try it.”

> College tutor 5:

> “..if that makes [trainees] happy, then, well, so be it because we all need to treat them as adult learners..So you know, your model might even make it easier..might suit us [better]. Who knows?”

The interview discussions revealed a strong appetite from the trainers to get the ball rolling and start a regional teaching programme as soon as possible. There was broad agreement that any initial programme would be a starting point and they were open to making changes along the way toward creating a robust programme that would stand the test of time and give high trainee satisfaction. All college tutors expressed support for the programme and agreed to take responsibility for any hospital-based teaching sessions initially to help get a programme off the ground. All trainees interviewed thought a programme would be a good idea and gave suggestions about how to collect feedback and ensure continuity of a programme and keep its high standards.

> College tutor 1:

> “I think the first thing you can, we can all say in this is there’s a need. What the need is I think is a real challenge but, and then also please go easy on yourself because you’ll not create the thing that is required here. And I think you know, putting something on paper, creating a structure, getting people in the same place, and allowing this to evolve into what it becomes is probably, I would argue, the most important thing. And so you’ve already achieved that by getting this happening.”

> School board member 3

> “The best thing to do is carry on the program and evaluate it on a 3 monthly basis or quarterly.”

## Discussion

Trainers and trainees were in favour of developing a regional teaching programme for Stage 1 anaesthetists in the deanery. Such a programme was seen to be a solution for some of the challenges faced by both trainers and trainees in navigating a new, demanding curriculum for Stage 1 trainees who work in very different hospital sites across our deanery.

Exam-based and extracurricular teaching, clinical experience, departmental and hospital ‘culture’ are some of the many ingredients required to build a strong Stage 1 training programme. Our findings demonstrated how challenging it can be to deliver a fair deanery wide programme that can help prepare Stage 1 trainees for their career ahead.

Our findings emphasised that teaching is an undervalued activity. Trainers are significantly undercompensated for the time, travel and effort required to prepare and execute a good session. Differing policies in compensation and time across hospitals limit the ability of educators to collaborate and innovate. Trainees’ clinical and service provision responsibilities are frequently placed ahead of their educational needs, and they too are poorly compensated for their time and travel. Too often, trainees are asked to take responsibility for their own teaching, leading to more variability in experience and potential gaps in their knowledge. Existing tools such as the GMC survey [1] are often used as rough measures of quality but provide insufficient detail to enable decision-makers to properly understand problems and develop solutions at a deanery level.

This study demonstrated the utility of using qualitative methods to answer key questions in the Model for Improvement [9] for a quality improvement project. The data provided a rich insight into trainers and trainees’ thoughts on the issues and allowed clearer characterisation of what we are trying to accomplish, how we know if change is an improvement, and what changes can be made that will result in improvement before implementation of any changes. The process of qualitative interviews also improved stakeholder engagement – interviewees appreciated the opportunity to share their thoughts on regional teaching and stage 1 anaesthetic training and were interested in working toward making their ideas a reality.

Our findings have directly led to specific actions for our QIP, including the design of a specific regional teaching survey for all stage 1 trainees, enquiries to the deanery and individual directors of medical education to understand policies for teaching and study budgets and leave, and engagement of consultants interested in leading Stage 1 teaching in each hospital.

This is the first study that the authors are aware of that explores perceptions of regional teaching in the United Kingdom, which we hope will stimulate further work and discussion about regional teaching programmes from other specialties and deaneries.

A limitation of this study was the small number of trainees interviewed, which may have introduced selection bias of trainees who were keen on discussing education and their training and missed some viewpoints from other trainees. We have addressed this by setting up a mandatory survey for Stage 1 trainees within our deanery with white-spaced questions to ensure that all trainees can share their views.

Another limitation is bias introduced by Author1’s role as an interviewer. Author1 is a senior registrar in the deanery, peer to the trainees and a junior to the consultants interviewed. It is possible that participants were reluctant to express their real thoughts, or left gaps in their narratives during interviews. The frankness displayed in the interviews when discussing topics such as funding, poor experiences and disparities between centres appear to suggest that Author1’s presence caused minimal disruption to the data.

Stage 1 is a new and complex period of training for anaesthetists. The rotational model of training can introduce significant disparities in trainee and trainer experience of Stage 1. A well-executed regional teaching programme may solve some of these problems but requires engagement and investment from trainers and trainees to succeed and sustain its quality.

## Supporting information

COREQ checklist

## Data Availability

All data produced in the present study are available upon reasonable request to the authors

## Acknowledgements

AL designed and led the project, performed the interviews and data collection, drafted the manuscript and edited the manuscript based on feedback from other authors.

RC, SJ, MR, SS and RW transcribed and anonymised the data, analysed the data using thematic analysis, reviewed the manuscript for important intellectual content and gave final approval for the submitted manuscript.

All authors agree to be accountable for all aspects of the work in ensuring that questions related to the accuracy or integrity of any part of the work are appropriately investigated and resolved.

The authors would like to acknowledge the following colleagues who helped with the transcription of the manuscripts and data coding:

Dr Roland Amoah

Dr Jiali Gao

Dr Farid Garas

Dr Ayushi Pandey

Dr Omar Risk

Dr Frances Rose

## COREQ supplementary information

AL is a female DPhil candidate at the University of Oxford and an anaesthetic registrar doctor (ST6) in Thames Valley. She was employed as a Trainee Improvement Fellow by Health Education England Thames Valley to complete this work.

Prior to conducting the study, she was awarded a postgraduate diploma (PGDip) in healthcare research methods (Merit) by the University of Oxford. During her PGDip, she completed a course on introduction of qualitative methodology by the Department of Continuing Education at the University of Oxford.

## Ethics and study registration

A detailed project proposal was submitted to the University of Oxford’s study classification group (consisting of Research Governance, Ethics & Assurance and R&D team members) and classified as service evaluation in December 2022.

Ethical approval is not necessary for service evaluation studies.

The study was registered as a trainee improvement project with Health Education Thames Valley.

A completed Consolidated Criteria for Reporting Qualitative Research (COREQ) checklist [8] is provided in the appendix.

